# The effect of education level between PIR and myopia: An interaction and mediation analysis

**DOI:** 10.1101/2025.03.18.25324210

**Authors:** Jiaqi Chen, Cong Zhao, Tongtong Chen, Zheyu Bao

**Affiliations:** Eye School of Chengdu University of Traditional Chinese Medicine, Sichuan Province, China; Key Laboratory of Sichuan Province Ophthalmopathy Prevention & Cure and Visual Function Protection with TCM Laboratory, Sichuan Province, China; Liaoning University of Traditional Chinese Medicine China

**Author notes:** Corresponding author at: Chengdu University of Traditional Chinese Medicine, Sichuan Province, China.

## Abstract

**Background:** The role of socioeconomic factors, particularly educational attainment, in the relationship between income and myopia remains unclear. As the poverty income ratio (PIR) reflects economic status, understanding its interaction with education on myopia prevalence is essential for informing public health and clinical strategies.

**Methods:** This cross-sectional study analyzed NHANES data (1999–2008) with 28,867 participants. Weighted multivariable logistic regression, generalized additive models, and mediation analysis were used to assess PIR’s association with myopia and the mediating/moderating role of education.

**Results:** PIR was significantly associated with myopia prevalence (OR = 1.05, 95% CI: 1.02– 1.08), but the association varied by age, gender, and education.Age: PIR was positively associated with myopia in individuals <60 years, but not in those ≥60.Gender: No significant association in males, while in females, PIR was positively linked to myopia (Right eye: OR = 1.05; Left eye: OR = 1.07).Education: In lower-education groups (Grade 9–11 and below), PIR and left-eye myopia showed a threshold effect at PIR = 2.9 (PIR < 2.9: OR = 0.92; PIR > 2.9: OR = 1.08). The right eye showed a similar but nonsignificant trend. In higher-education groups, PIR exhibited a linear association with left-eye myopia (OR = 1.08) and a nonlinear association with right-eye myopia, with risk increasing sharply at PIR = 4.44 (OR from 1.05 to 1.29). Education mediated 20% of PIR’s effect on right-eye myopia and 24% on left-eye myopia.

**Conclusion:** Education plays a crucial mediating and moderating role in the PIR-myopia relationship, with a stronger effect in the left eye. The findings suggest integrating socioeconomic and educational factors into myopia prevention strategies to identify high-risk populations.

## Introduction

Myopia is a common visual disorder and has become a significant global public health concern, with its prevalence steadily increasing in recent years[1,2]. As one of the leading causes of visual impairment, myopia not only affects quality of life but also imposes a substantial economic burden on individuals and healthcare systems[3]. The etiology of myopia is multifactorial, involving both genetic and environmental factors[4,5]. Among these, socioeconomic factors have increasingly been recognized as potential contributors to the prevalence of myopia, with the poverty-income ratio (PIR) serving as a quantitative indicator of household economic status. However, the association between economic and educational factors and myopia prevalence may vary across populations, necessitating further investigation.

Education is a well-established risk factor for myopia, with numerous studies indicating that higher educational attainment increases the risk of developing myopia[6,7]. Individuals from high-income families may have better access to educational resources, leading to prolonged study time and increased near-work exposure—both of which are strongly associated with myopia progression[8]. However, individuals from low-income backgrounds may experience different nutritional conditions[9,10], educational levels, and lifestyle habits[11,12], potentially contributing to population differences in the relationship between PIR and myopia prevalence. It is crucial to explore whether education mediates the association between PIR and myopia prevalence and to determine the extent of this mediation.

Additionally, previous epidemiological studies on myopia have often used data from a single eye to represent the relationship between refractive status and various variables, overlooking ocular laterality—the inherent differences in refractive status between the left and right eyes[13]. Some studies suggest that associations between unilateral or bilateral visual impairment and physiological indicators may differ and may also vary between males and females[14]. When examining the role of education in the association between PIR and myopia, it is essential to consider ocular laterality, as potential hemispheric functional asymmetry may differentially affect myopia in each eye. Ignoring ocular laterality could lead to biased estimates of the relationship among PIR, education, and myopia risk. Therefore, this study will analyze the left and right eyes separately to investigate whether PIR and its interaction with education have distinct associations with myopia prevalence in each eye.

This study aims to examine the association between PIR and myopia prevalence in both eyes, the mediating role of education in this relationship, and potential subgroup differences. Our findings may provide novel insights into the socioeconomic determinants of myopia and offer support for more targeted public health interventions.

## 2. Method

### 2.1 Study populations

We utilized data from the 1999–2008 National Health and Nutrition Examination Survey (NHANES), a cross-sectional series of interviews and examinations conducted by the Centers for Disease Control and Prevention (CDC) to assess the health and nutritional status of the non-institutionalized civilian population in the United States. NHANES aims to provide nationally representative health statistics, and its de-identified data are publicly accessible. To ensure accurate estimations of disease prevalence in the U.S. population, NHANES employs a complex, stratified, multistage probability sampling design with appropriate weighting adjustments.

The study protocol received approval from the Ethics Review Board of the National Center for Health Statistics (NCHS). Written informed consent was obtained from all participants at the time of recruitment. For this analysis, we excluded 20,349 individuals with missing spherical equivalent measurements for either the left or right eye, as well as 2,384 participants lacking data on the poverty-income ratio (PIR). Additionally, 23 cases were removed due to incomplete covariate information. Consequently, the final analytic sample consisted of 28,867 participants. A detailed flowchart illustrating the sample selection process is provided in the corresponding figure.

### 2.2 Survey variables

In this study, the primary exposure variable was the poverty-income ratio (PIR), while the outcome variable was the presence or absence of myopia among participants. All NHANES participants aged 12 years and older were eligible for refractive examinations, except for those with blindness or severe ocular infections. The assessment of refractive status was conducted using the Nidek Auto Lensmeter Model LM-990A and the Nidek Auto Refractor Model ARK-760, which measured the refractive components of the eye, including sphere, cylinder, and axis. An objective evaluation of refractive error was performed without cycloplegia. Three consecutive measurements of both spherical and cylindrical power were recorded, and the mean value was calculated. The spherical equivalent (SphEq) was determined by adding half of the cylindrical power to the spherical power. In this study, myopia was defined as an SphEq of ≤ −0.75 diopters.

The PIR was calculated by dividing household income by the federal poverty threshold and was further adjusted for inflation using the Consumer Price Index (CPI) to derive a standardized income index. Based on PIR values, participants were categorized into three income groups: low-income (PIR ≤ 1.3), middle-income (PIR between 1.3 and 3.5), and high-income (PIR > 3.5).

Additionally, this study accounted for several covariates, including age, sex, race/ethnicity, educational attainment, smoking status, diabetes, body mass index (BMI), and serum vitamin D3 levels. Detailed definitions and descriptions of all variables used in the study are available on the official NHANES website (https://www.cdc.gov/nchs/NHANES).

### 2.3 Statistical analysis

All statistical analyses were conducted in accordance with the data analysis guidelines provided by the CDC, employing appropriate NHANES sampling weights to account for the survey’s complex, multistage, clustered sampling design. Descriptive analyses were performed to examine differences among participants across different PIR categories. For continuous variables, weighted t-tests were utilized, while weighted chi-square tests were applied for categorical variables. Continuous data were summarized as means with standard errors (SE), whereas categorical data were expressed as proportions.

To investigate the relationship between PIR and cataract prevalence, we implemented three distinct multivariable regression models, all of which incorporated NHANES sampling weights to account for the survey’s complex design. Model 1 was unadjusted for covariates, while Model 2 accounted for sex, age, and race. Model 3 was further adjusted for smoking status, educational attainment, body mass index (BMI), and diabetes status.

In the sensitivity analysis, PIR was categorized into three groups—high, middle, and low-income levels—and a trend test was conducted to assess the robustness of the results.Additionally, generalized additive models (GAM) with smooth curve fitting were employed to explore the association between PIR and the prevalence of myopia. In instances where a nonlinear relationship was detected, a two-segment linear regression model (also referred to as a segmented regression model) was applied to delineate interval-specific associations and assess potential threshold effects. The presence of a threshold effect was determined using a likelihood ratio test, comparing the fit of a single-piece (non-segmented) model with that of a two-segment model. The breakpoint (K) in the segmented model was estimated using the maximum likelihood method and refined through a two-step recursive approach.

Subgroup analyses were conducted using stratified multivariable logistic regression models, considering stratification factors such as sex (male/female), age groups (<18, 18–40, 40–60, ≥60 years), Serum vitamin D3 status (deficient/insufficient/sufficient), and BMI categories (normal weight/overweight). Furthermore, interaction terms incorporating these stratification factors were included as predefined potential effect modifiers, with likelihood ratio tests used to evaluate heterogeneity in associations across subgroups. In addition, smooth curve fitting was utilized to further investigate the potential linear or nonlinear relationship between PIR and myopia prevalence.

Missing data were addressed using median imputation for continuous variables and mode imputation for categorical variables. All statistical analyses were performed using R software (version 4.1.3, The R Foundation) and Empower software (X&Y Solutions, Inc., Boston, USA). A two-sided p-value <0.05 was considered statistically significant.

## 3. result

### 3.1 Study population

The baseline characteristics of the study population (**Table 1**) indicate that a total of 28,867 participants were included in the analysis, comprising 14,097 males (45.14%) and 14,770 females (51.17%). Among them, 13,110 individuals (48.4%) were diagnosed with myopia in the right eye, while 13,031 participants (45.14%) had myopia in the left eye. The age of participants ranged from 12 to 85 years, with a mean age of 37.55 years. The average PIR was recorded at 2.47 (SD = 1.6).

In terms of income distribution, 9,292 participants were classified as low-income, 10,924 as middle-income, and 8,651 as high-income. Notably, although individuals in the high-income group had better financial status and higher levels of vitamin D3, they also exhibited the highest prevalence of myopia in both eyes. Compared to those in the low-income group (PIR <1.3), individuals in the high-income category (PIR >3.5) tended to be older, had a lower prevalence of smoking, slightly higher BMI, a reduced prevalence of diabetes, and a greater level of educational attainment. Additionally, their Serum vitamin D3 concentrations were notably higher. Moreover, a greater proportion of individuals in the high-income group were identified as non-Hispanic White.

### 3.2 The relationship between PIR and myopia

We conducted multivariable linear regression analyses using three models to examine the relationship between PIR and myopia prevalence, as shown in **Table 2**. Our findings reveal that PIR is positively associated with the prevalence of myopia. A significant positive association between PIR and the prevalence of myopia in both eyes was observed in both the unadjusted model and the minimally/fully adjusted models. In the fully adjusted model, for every 1-unit increase in PIR, the prevalence of myopia in the right eye increased by 3% (Model 3: OR = 1.03, 95% CI: 1.01, 1.05), and the prevalence in the left eye increased by 3% (Model 3: OR = 1.03, 95% CI: 1.01, 1.05). Even when PIR was categorized into low, medium, and high levels, participants with the highest PIR (PIR > 3.5) had a 9% higher prevalence of myopia in the right eye and an 11% higher prevalence in the left eye compared to those with the lowest PIR (PIR ≤ 1.3), with a significant trend test. Additionally, we fitted a smooth curve to examine the relationship between PIR and myopia in both eyes. The results indicated a positive association between PIR and the prevalence of myopia in both eyes (Figure 2), suggesting that PIR is positively correlated with myopia in both eyes.

**Figure 1.**
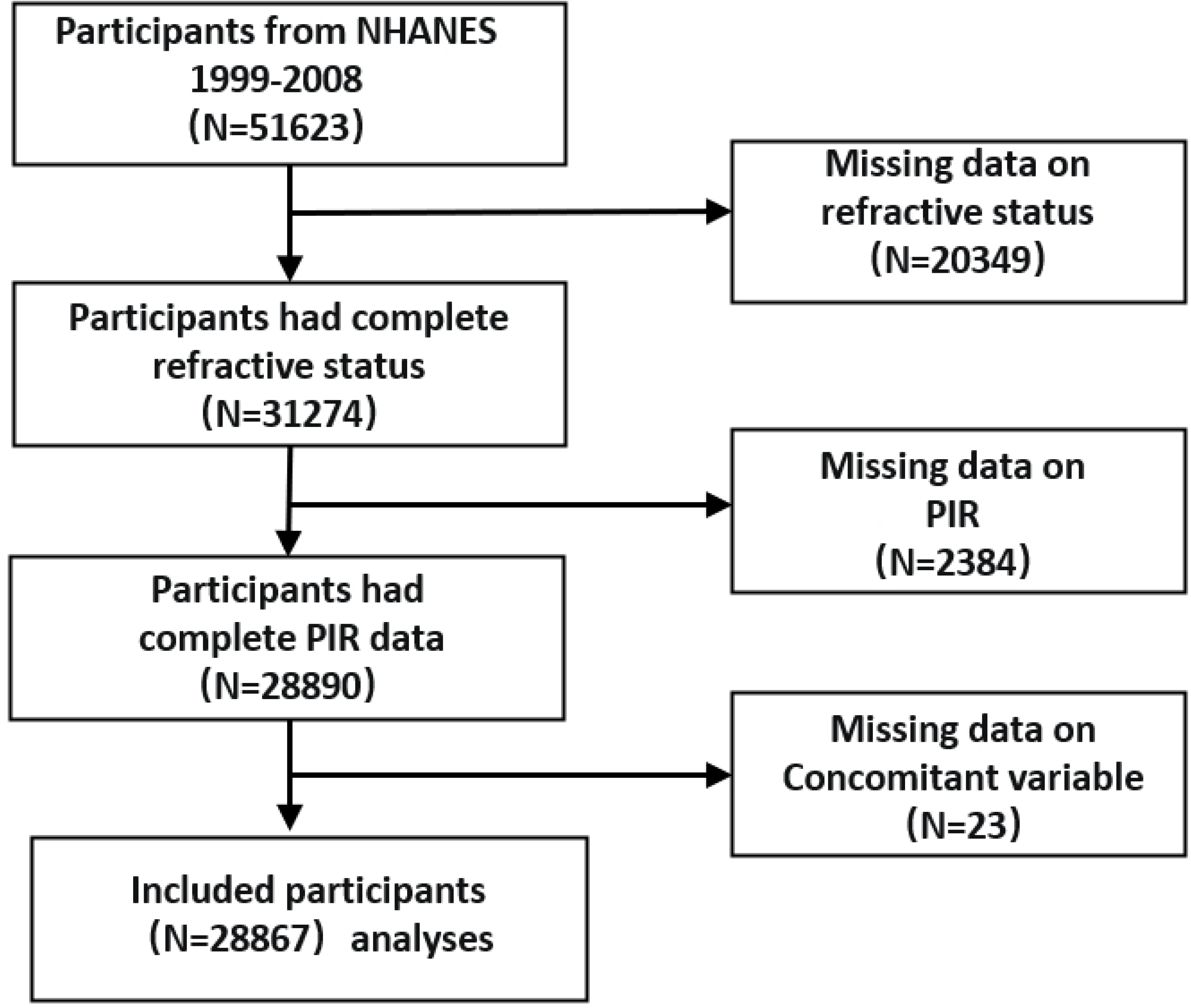
Flow chart of participant selection NHANES, National Health and Nutrition Examination Survey. PIR, family income to poverty ratio.

**Figure 2.**
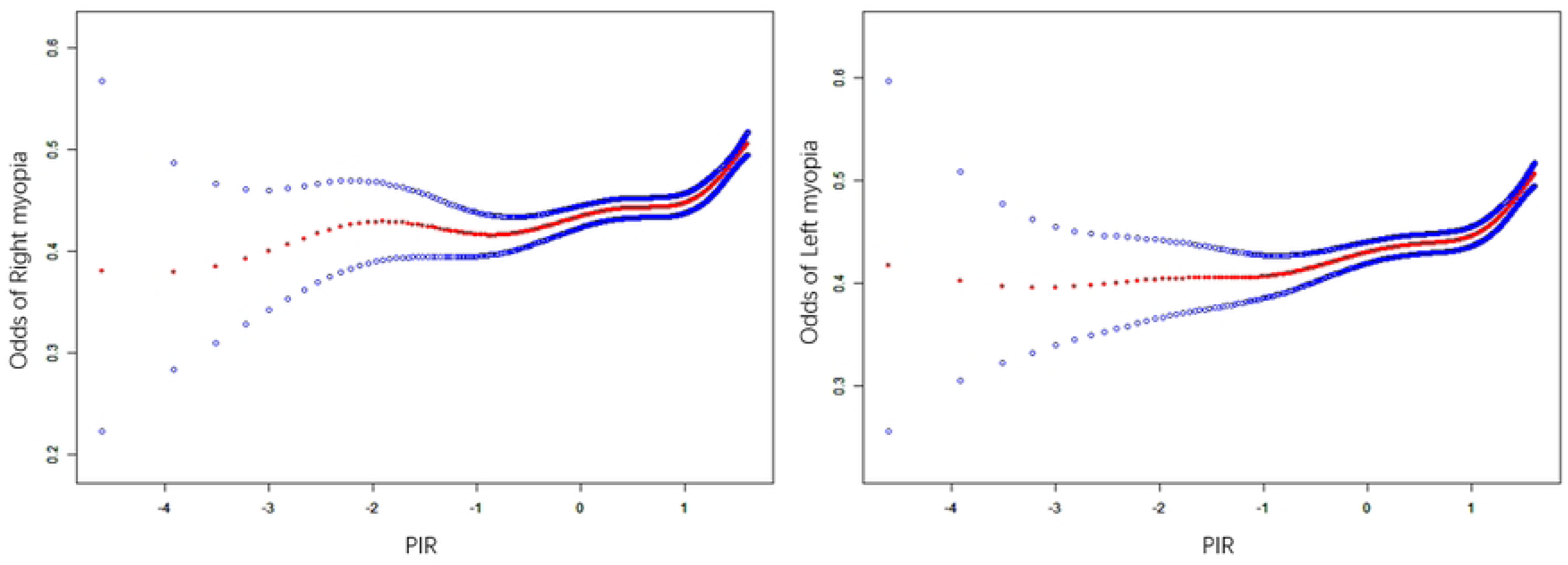
The relationship between PIR and myopia prevalence. The red solid line represents the smoothed fitted curve between the variables, while the blue shaded area indicates the 95% confidence interval of the fitted model. The analysis was adjusted for age, sex, race, body mass index, education level, smoking status, diabetes status, and serum vitamin D3 levels.

#### Subgroup Analysis and Interaction Test

To further assess the robustness of the association between PIR and myopia prevalence, we conducted subgroup analyses and interaction tests. **Table 3** presents the results across different stratified variables, including sex, race, BMI, smoking status, diabetes status, education level, and serum vitamin D3 concentration. Our findings indicate that race, smoking status, BMI, diabetes status, and serum vitamin D3 levels did not significantly modify the positive association between PIR and myopia prevalence. However, notable variations were observed across different age groups, sexes, and education levels, with a consistent pattern observed in both eyes.

Specifically, in the male subgroup, PIR was not significantly associated with myopia prevalence in either eye. In the age-stratified analysis, participants were categorized into four groups: adolescents (<18 years), young adults (18-40 years), middle-aged adults (40-60 years), and older adults (>60 years). The results showed a positive correlation between PIR and myopia prevalence in both eyes among individuals younger than 60 years, whereas no statistically significant association was detected in those aged 60 years and above.

When education level was examined as an effect modifier, it was found that among individuals with lower educational attainment (Less Than 9th Grade, 9-11th Grade), PIR was not significantly associated with myopia prevalence in either eye. However, in individuals with moderate to high educational levels (High School Graduate, Some College or AA degree, College Graduate or above), a significant positive association was observed between PIR and myopia prevalence in both eyes, with a stronger effect in the left eye (left eye OR = 1.08 > right eye OR = 1.06). Further analysis revealed that among highly educated individuals (College Graduate or above), each 1-unit increase in PIR was associated with a 6% increase in myopia risk in the right eye (95% CI: 1.02-1.10) and an 8% increase in the left eye (95% CI: 1.03-1.12).

### 3.4 Threshold Effect Analysis of the Association Between PIR and Myopia Across Different Educational Levels Using a Two-Segment Linear Regression Model

To explore the nonlinear relationship between PIR and myopia prevalence, we employed Generalized Additive Models (GAM) and smoothed curve fitting. Based on the results of interaction tests and clinical relevance, education level was categorized into two groups (Grade 9-11th and below / High School Graduate and above) for threshold effect analysis, as illustrated in **Figure 3** and **Table 4**.

**Figure 3.**
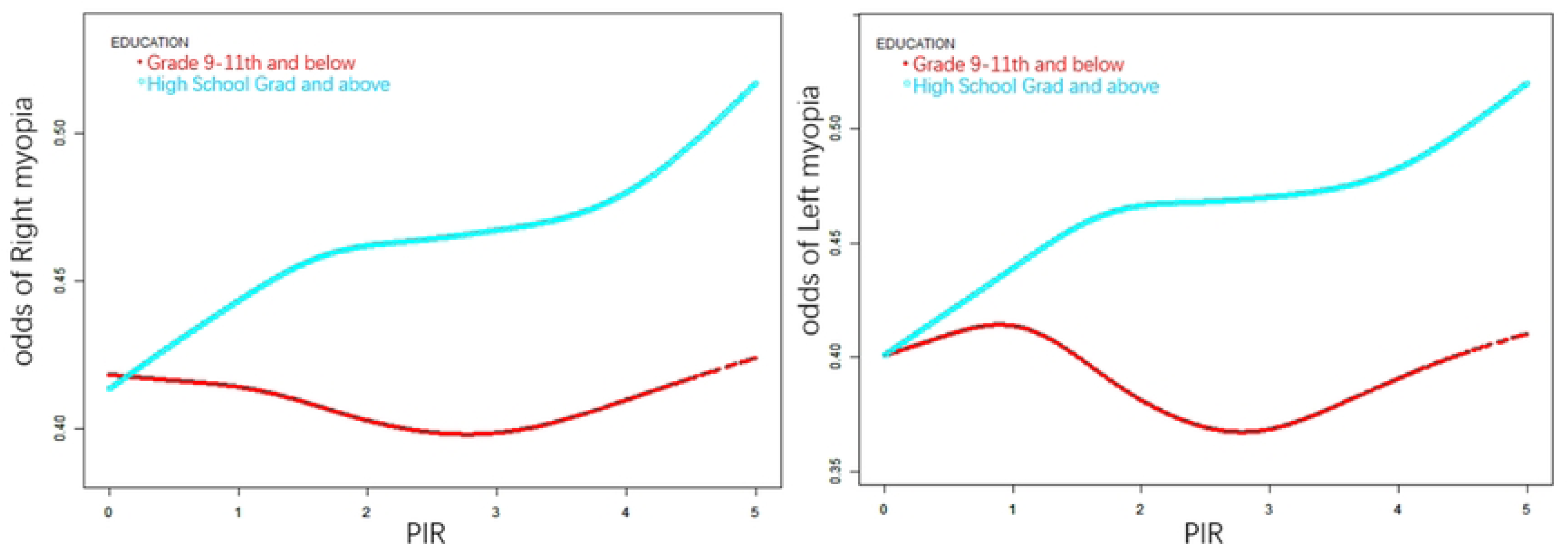
When education level was categorized into two groups (Grade 9-11th and below / High School Graduate and above) as an effect modifier, the relationship between PIR and myopia prevalence was analyzed. The red solid line represents the smoothed curve fitting for individuals with an education level of Grade 9-11th and below, while the blue band represents the smoothed curve fitting for those with a High School Graduate education level and above. The analysis was adjusted for age, sex, race, body mass index (BMI), education level, smoking status, diabetes status, and serum vitamin D3 levels.

Our findings indicate that in the Grade 9-11th and below group, PIR was not significantly associated with myopia prevalence in the right eye. However, among individuals with a High School Graduate education level and above, a nonlinear relationship was observed between PIR and right-eye myopia prevalence. Through a two-segment linear regression model, we identified a breakpoint (K) at 4.44. On both sides of this threshold, PIR exhibited a positive association with right-eye myopia prevalence. However, the effect size varied: on the left side of the breakpoint, the positive association was relatively weaker (OR = 1.05, 95% CI: 1.02-1.08), with each 1-unit increase in PIR corresponding to a 5% increase in myopia prevalence. In contrast, on the right side of the breakpoint, the association was considerably stronger (OR = 1.29, 95% CI: 1.10-1.52), where each 1-unit increase in PIR led to a 29% rise in myopia prevalence.

As shown in **Table 5**, for the left eye, the association between PIR and myopia prevalence among individuals with a High School Graduate education level and above followed a linear positive trend (OR = 1.08, 95% CI: 1.06-1.10), indicating that each 1-unit increase in PIR corresponded to an 8% rise in myopia prevalence. In contrast, in the Grade 9-11th and below group, a nonlinear association was observed. The breakpoint (K) for this group was identified at 2.9. To the left of this threshold, PIR was negatively associated with myopia prevalence (OR = 0.92, 95% CI: 0.87-0.98), suggesting that each 1-unit increase in PIR was linked to an 8% reduction in myopia prevalence in the left eye. However, on the right side of the breakpoint, the association shifted to a linear positive correlation (OR = 1.08, 95% CI: 1.06-1.10), where each additional unit of PIR was associated with an 8% increase in myopia prevalence.

### 3.5. Mediation effect analysis

We further employed mediation analysis to investigate the mediating role of education level in the relationship between PIR and myopia prevalence. Education level exhibited a significant indirect effect (mediating effect) on both left and right eye myopia prevalence, with mediation rates of 20% for the right eye and 24% for the left eye (Figure 4, Tables 6-7).

**Figure4.**
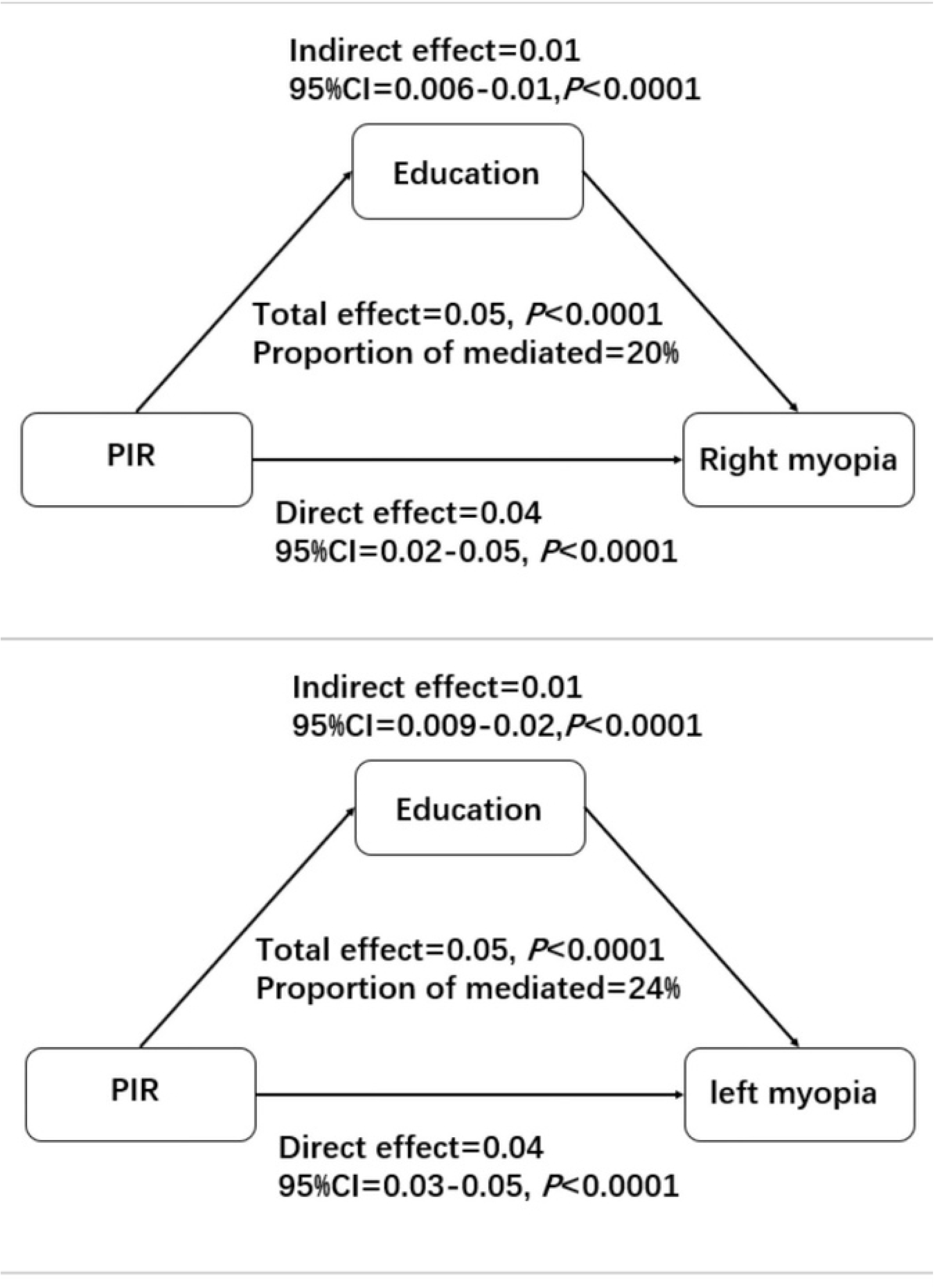
Illustration of the Mediation Proportion of Educational Level in the Association Between PIR and Myopia Prevalence

## 4. Discussion

This study is the first to reveal the mediating role of educational attainment in the association between PIR and myopia prevalence, while also identifying an eye-specific difference, with the mediation proportion being greater for the left eye than for the right eye. Furthermore, when stratified by educational level, distinct population differences were observed. Among individuals with lower educational attainment (Grade 9-11th and below), the relationship between PIR and left-eye myopia prevalence exhibited a nonlinear pattern. A threshold effect was identified at a breakpoint (K) of 2.9. To the left of this breakpoint, PIR was negatively associated with left-eye myopia prevalence (OR = 0.92, 95% CI: 0.87–0.98), indicating that for every one-unit increase in PIR, the prevalence of left-eye myopia decreased by 8%. However, to the right of this breakpoint, the association shifted to a linear positive correlation (OR = 1.08, 95% CI: 1.06–1.10). Although the association for the right eye did not reach statistical significance, the fitted curve suggested a similar trend.In contrast, among individuals with higher educational attainment (High School Grad and above), PIR exhibited a nonlinear positive correlation with right-eye myopia prevalence. A breakpoint (K) was identified at 4.44, with a more pronounced effect beyond this threshold. Specifically, to the left of the breakpoint, each unit increase in PIR was associated with a 5% increase in right-eye myopia prevalence, whereas to the right of the breakpoint, the effect was considerably stronger, with each unit increase in PIR leading to a 29% rise in myopia prevalence. For the left eye, the association was a consistent linear positive correlation, with every unit increase in PIR corresponding to an 8% increase in left-eye myopia prevalence.Overall, despite minor differences between the two eyes, a general pattern emerged: in individuals with higher educational attainment (High School Grad and above), PIR was positively associated with myopia prevalence in both eyes. In contrast, among those with lower educational attainment (Grade 9-11th and below), the relationship between PIR and myopia prevalence followed a U-shaped nonlinear pattern, with a breakpoint (K) identified at 2.9.

Numerous studies have indicated a correlation between PIR and the prevalence of myopia. For example, a cross-sectional study in the United States involving 6,814 participants aged 12 to 19 years found a positive association between higher levels of PIR and a greater incidence of high myopia[15]. Additionally, while there are regional and national variations, a consistent positive correlation between socioeconomic status and myopia prevalence has been observed globally. For instance, a cross-sectional study in South Korea, which included 8,633 participants under the age of 19, identified monthly household income as a significant determinant of myopia prevalence[16]. The study also found that participants living in parent-owned homes or urban areas were more likely to develop myopia. This suggests that, whether in Asian or European populations, higher family income tends to correlate with a higher prevalence of myopia^2^.These findings highlight the broader pattern that economic factors, particularly family income, are significant contributors to the prevalence of myopia.However, some studies have reported contrasting findings[19], which may be attributed to differences in the study populations. Based on the results of this study, it can be explained that in lower education groups, there is a U-shaped relationship between PIR and myopia prevalence. Specifically, lower PIR levels are associated with a higher likelihood of factors such as nutritional deficiencies or uncorrected refractive errors, leading to a negative correlation between PIR and myopia prevalence. When PIR exceeds the Breakpoint (K), however, extended education duration and increased indoor activities contribute to a shift toward a positive correlation between PIR and myopia prevalence.Additionally, there exists a paradox in the high myopia rates across different regions. While studies from various countries and regions have confirmed a positive correlation between income levels and myopia prevalence, many high-income developed nations exhibit lower myopia rates compared to some developing countries[20]. This discrepancy may be attributed to the mediating role of educational pressure, which differs due to cultural factors. In countries influenced by Confucianism, such as China and South Korea, education is viewed as a primary means of social mobility, and families may be more inclined to sacrifice other resources in order to invest in education, thereby amplifying the relationship between PIR and myopia[21].This study found that educational level mediated 20-24% of the relationship between PIR and myopia prevalence, offering a compelling explanation for this phenomenon. The observed differences in the mediation effect between the left and right eyes (with the left eye showing a higher mediation effect) can be explained through the following reasoning.First, the differences in the mediation effects between the left and right eyes may be attributed to the lateralization of brain function influenced by education. The left eye is associated with the visual cortex in the right hemisphere of the brain, which is more active during spatial tasks such as geometry and chart analysis[22,23]. Intense educational activities could enhance the training of these tasks, leading to more pronounced activation of the right hemisphere. Karlsson (2008) proposed that myopia-related genes may simultaneously impact both brain development and eye growth. These genes may regulate the axial length of the eye while promoting neural development, indicating genetic pleiotropy[24]. Additionally, a study on children in Singapore suggested that brain development (e.g., the size of the neocortex) and eye development could be regulated by the same biological pathways. For example, growth factors or neuro-signals may concurrently affect intelligence and myopia[25]. If the hypothesis of a shared developmental pathway between myopia and cognitive abilities holds true, the increased activity of the right hemisphere could correspond to accelerated axial elongation in the left eye.However, this remains a core hypothesis of the myopia-cognition association, and specific molecular pathways require further investigation. Moreover, the differences in eye regulation functions can also explain this. During reading or learning tasks, the non-dominant eye (left eye) may excessively compensate by over-regulating, which exacerbates myopia development as the demand for visual accommodation increases. This tilting behavior leads to greater convergence angles and shorter reading distances for the left eye in the majority of right-handed individuals, intensifying accommodation fatigue and lag[26]. Despite similar myopia prevalence in both eyes within the overall population, when refractive differences occur, the right eye may even have a higher prevalence than the left[27,28]. However, the mediation effect of education level suggests that the left eye is more sensitive to the influence of education, indicating that the right eye’s myopia progression may be more dominated by genetic or early developmental factors, while the left eye’s progression is more influenced by educational mediators. This reflects the eye-specific heterogeneity due to later behavioral impacts. In terms of exploring the mechanisms, it is important to quantify the differences in fixation time and accommodative fluctuations between the left and right eyes during education-related tasks.

This study also identified gender differences in the relationship between PIR and myopia, which may result from a combination of factors including environmental exposures (outdoor activity, educational investment), biological susceptibility (hormones, metabolism), and sociocultural constraints (health management, gender roles). First, in terms of lifestyle, women in higher PIR households may prefer indoor, quiet activities that require close-up vision, while men are more likely to engage in outdoor sports[29],[30]. Furthermore, hormonal differences may also play a role. Estrogen promotes the onset and progression of myopia by regulating corneal thickness, scleral remodeling, and gene expression, while insufficient testosterone weakens the myopia-inhibiting effects of retinal dopamine[31,32]. Additionally, women in low-income households may be more prone to cortisol-melatonin rhythm disturbances[33], and prolonged elevated cortisol levels, through activation of the HPA axis (hypothalamic-pituitary-adrenal axis), inhibit the hypothalamus from secreting gonadotropin-releasing hormone (GnRH), thereby reducing the pituitary secretion of follicle-stimulating hormone (FSH) and luteinizing hormone (LH), ultimately decreasing ovarian estrogen synthesis[34]. As a result, low-income women may experience a delayed impact of estrogen on myopia compared to high-income women with higher estrogen levels. More potential mechanisms require further investigation.

Additionally, this study revealed age-related differences in the association between PIR and myopia. Specifically, the relationship between myopia prevalence and PIR is stronger in individuals under 60 years of age, while no significant relationship is found in individuals aged 60 and older. This may be due to the fact that myopia predominantly develops in earlier life stages. Factors such as PIR (which includes education pressure and nutrition levels) have a greater impact on younger populations. For individuals over 60, the demand for high-intensity near-vision activities and education pressure significantly decreases compared to younger populations, and the likelihood of indoor work and screen time is lower among older adults[35–37].Preventing and controlling myopia requires a comprehensive approach that integrates educational factors, rather than relying solely on income levels. However, the mediating role of education remains limited. The influence of PIR-related factors on myopia is still an area worth exploring. Given the complex, nonlinear nature of the relationship between PIR and myopia, clinical interventions can be informed to better target high-risk myopia populations. More precise identification of individuals, especially women under 60 with high household income and moderate to high education levels, is essential for focusing on myopia progression. Policy interventions should aim to alleviate educational pressures to reduce their impact on myopia development.

This study has several strengths. First, it is based on a nationally representative dataset, enhancing the generalizability of the findings. Additionally, multiple covariates were adjusted in the regression analysis to ensure a more accurate interpretation of the observed associations. The large sample size also allowed for subgroup analyses, further validating the robustness of the results. More importantly, this study highlights the role of economic factors in the clinical management of myopia and public policy formulation—an aspect often overlooked—while providing valuable insights for practical applications. Lastly, our findings offer some support for the hypothesis of cerebral hemispheric lateralization in ocular development influenced by educational factors, providing a potential reference for future mechanistic research.

Nevertheless, it is important to acknowledge the limitations of this study. Although we have adjusted for several key covariates, numerous unaccounted factors, such as occupational type, living environment, and psychological influences, may still contribute to the prevalence of myopia. Additionally, the Serum vitamin D3 levels included in this study may not fully capture the participants’ actual outdoor light exposure. To ensure a sufficiently large sample size, the database lacked comprehensive data on outdoor activity, preventing a more detailed statistical analysis. These unmeasured confounding factors may affect the accuracy of our findings. Furthermore, as the data for this study were derived from a single country and focused on a specific population, the generalizability of our results to other racial, ethnic, or national groups remains uncertain. Most importantly, given the cross-sectional design of this study, we cannot infer causality or thoroughly explore the underlying mechanisms of the observed associations. Therefore, future longitudinal studies are needed to establish causal relationships and further investigate the impact of socioeconomic factors on the development of myopia.

## Data Availability

We utilized data from NHANES, a cross-sectional series of interviews and examinations conducted by the Centers for Disease Control and Prevention (CDC) on the civilian, non-institutionalized U.S. population. The purpose of NHANES is to provide national health statistics, and the de-identified data are publicly available. NHANES employs a stratified, multistage sampling design and weighting scheme to accurately estimate disease prevalence in the U.S. population. The study received approval from the National Center for Health Statistics (NCHS) Ethics Review Board, and all participants provided written informed consent at the time of recruitment.

## Supplementary Material

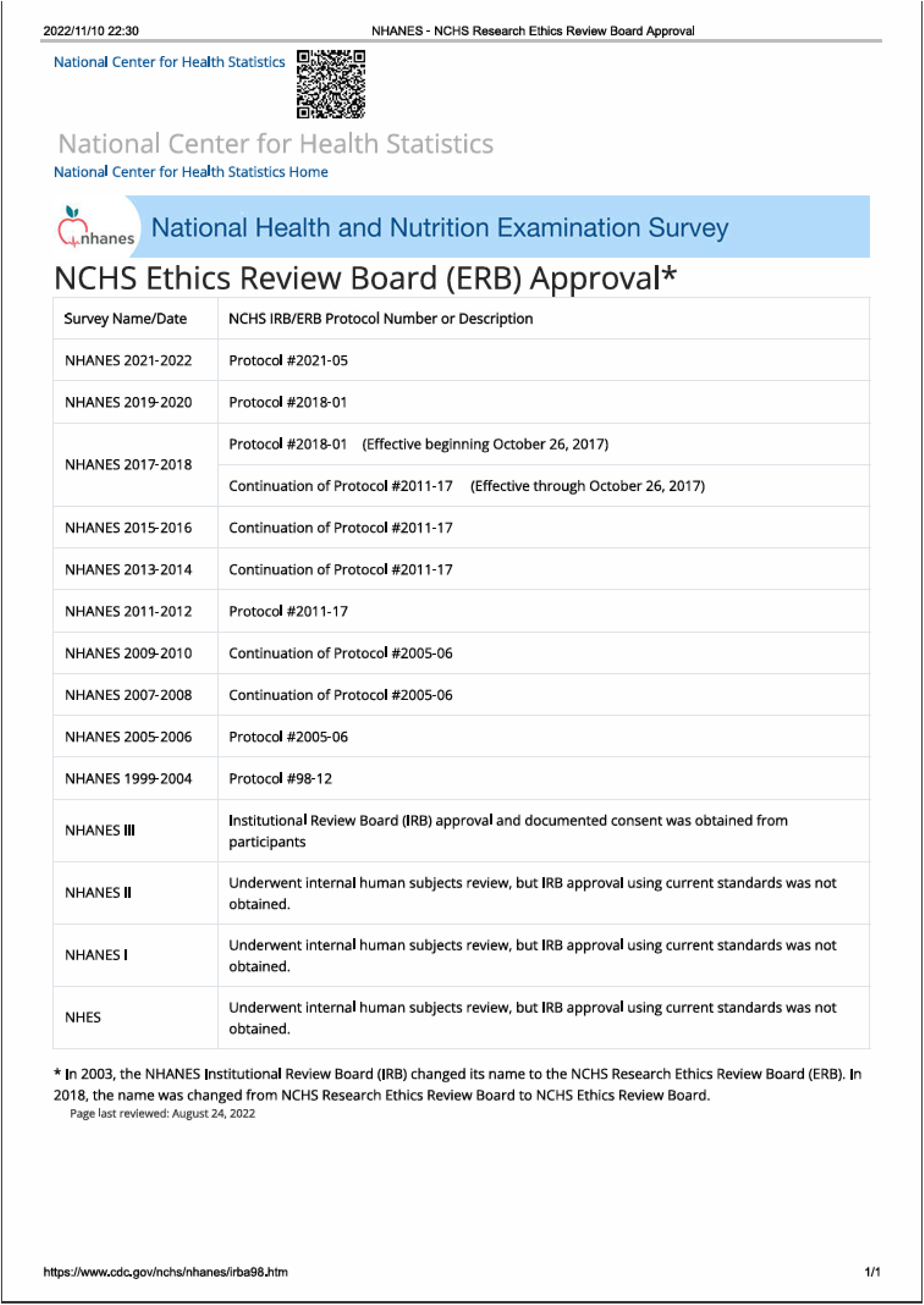

This study involved participants from 1999 to 2008

## References

1. George AS, George AH, Shahul A. The Myopia Epidemic: A Growing Public Health Crisis Impacting Children Worldwide. Partners Universal International Research Journal [Internet]. 2023 [cited 2025 Feb 21];2(3):120–38. Available from: https://www.puirj.com/index.php/research/article/view/136

2. Resnikoff S, Jonas JB, Friedman D, He M, Jong M, Nichols JJ, Ohno-Matsui K. et al. Myopia–a 21st century public health issue. Investigative ophthalmology & visual science [Internet]. 2019 [cited 2025 Feb 21];60(3):Mi–Mii. Available from: https://iovs.arvojournals.org/article.aspx?articleid=2727309

3. Pyne ALB, Noy A, Main KHS, Velasco-Berrelleza V, Piperakis MM, Mitchenall LA. et al. Base-pair resolution analysis of the effect of supercoiling on DNA flexibility and major groove recognition by triplex-forming oligonucleotides. Nat Commun. 2021 Feb 16;12(1):1053.

4. Sankaridurg P, Tahhan N, Kandel H, Naduvilath T, Zou H, Frick KD. et al. IMI Impact of Myopia. Invest Ophthalmol Vis Sci. 2021 Apr 28;62(5):2.

5. Guggenheim JA, Williams C, UK Biobank Eye and Vision Consortium. Role of Educational Exposure in the Association Between Myopia and Birth Order. JAMA Ophthalmol. 2015 Dec;133(12):1408–14.

6. Shapira Y, Mimouni M, Machluf Y, Chaiter Y, Saab H, Mezer E. The Increasing Burden of Myopia in Israel among Young Adults over a Generation: Analysis of Predisposing Factors. Ophthalmology. 2019 Dec;126(12):1617–26.

7. Williams KM, Bertelsen G, Cumberland P, Wolfram C, Verhoeven VJM, Anastasopoulos E. et al. European Eye Epidemiology (E(3)) Consortium. Increasing Prevalence of Myopia in Europe and the Impact of Education. Ophthalmology. 2015 Jul;122(7):1489–97.

8. Zhang C, Li L, Jan C, Li X, Qu J. Association of School Education With Eyesight Among Children and Adolescents. JAMA Netw Open. 2022 Apr 1;5(4):e229545.

9. Ngadze RT, Linnemann AR, Fogliano V. Nexus of local context and technology in child complementary feeding. Global Food Security [Internet]. 2023 Sep 1 [cited 2025 Feb 22];38:100714. Available from: https://www.sciencedirect.com/science/article/pii/S2211912423000445

10. Olza J, Martínez de Victoria E, Aranceta-Bartrina J, González-Gross M, Ortega RM, Serra-Majem L. et al. Adequacy of Critical Nutrients Affecting the Quality of the Spanish Diet in the ANIBES Study. Nutrients [Internet]. 2019 Oct 1 [cited 2025 Feb 22];11(10):2328. Available from: https://www.ncbi.nlm.nih.gov/pmc/articles/PMC6835880/

11. Abdullah A, Doucouliagos H, Manning E. Does Education Reduce Income Inequality? A Meta-Regression Analysis. Journal of Economic Surveys [Internet]. 2015 [cited 2025 Feb 22];29(2):301–16. Available from: https://onlinelibrary.wiley.com/doi/abs/10.1111/joes.12056

12. Leme ACB, Baranowski T, Thompson D, Nicklas T, Philippi ST. Sustained impact of the “Healthy Habits, Healthy Girls – Brazil” school-based randomized controlled trial for adolescents living in low-income communities. Preventive Medicine Reports [Internet]. 2018 Jun 1 [cited 2025 Feb 22];10:346–52. Available from: https://www.sciencedirect.com/science/article/pii/S2211335518300688

13. Cheng CY, Yen MY, Lin HY, Hsia WW, Hsu WM. Association of Ocular Dominance and Anisometropic Myopia. Invest Ophthalmol Vis Sci [Internet]. 2004 Aug 1 [cited 2025 Feb 22];45(8):2856–60. Available from: https://iovs.arvojournals.org/article.aspx?articleid=2124019

14. Nitzan I, Shakarchy N, Megreli J, Akavian I, Derazne E, Afek A. et al. Body mass index and visual impairment in Israeli adolescents: A nationwide study. Pediatr Obes. 2024 Jan;19(1):e13083.

15. Tao Q, Chang Y, Day AS, Wu J, Wang X. Association between serum 25-hydroxyvitamin D level and myopia in children and adolescents: a cross-sectional study. Transl Pediatr. 2024 Feb 29;13(2):310–7.

16. Lim HT, Yoon JS, Hwang SS, Lee SY. Prevalence and associated sociodemographic factors of myopia in Korean children: the 2005 third Korea National Health and Nutrition Examination Survey (KNHANES III). Jpn J Ophthalmol. 2012 Jan;56(1):76–81.

17. Jan C, Xu R, Luo D, Xiong X, Song Y, Ma J, Stafford RS. Association of Visual Impairment With Economic Development Among Chinese Schoolchildren. JAMA Pediatr. 2019 Jul 1;173(7):e190914.

18. Sperduto RD, Seigel D, Roberts J, Rowland M. Prevalence of Myopia in the United States. Archives of Ophthalmology [Internet]. 1983 Mar 1 [cited 2025 Mar 10];101(3):405–7. Available from: 10.1001/archopht.1983.01040010405011

19. Philipp D, Vogel M, Brandt M, Rauscher FG, Hiemisch A, Wahl S. et al. The relationship between myopia and near work, time outdoors and socioeconomic status in children and adolescents. BMC Public Health [Internet]. 2022 Nov 10 [cited 2025 Mar 10];22(1):2058. Available from: https://bmcpublichealth.biomedcentral.com/articles/10.1186/s12889-022-14377-1

20. Morgan IG, French AN, Ashby RS, Guo X, Ding X, He M. et al. The epidemics of myopia: Aetiology and prevention. Prog Retin Eye Res. 2018 Jan;62:134–49.

21. Morgan IG, Rose KA. Myopia and international educational performance. Ophthalmic Physiol Opt. 2013 May;33(3):329–38.

22. Mustafa Nadeem Kirmani, Mukesh Kumar Garg, Preeti Sharma. Parietal & Occipital Lobe Syndromes: Neuropsychological Approach. Int J Indian Psychol [Internet]. 2016 Mar 25 [cited 2025 Mar 10];3(2). Available from: https://ijip.in/articles/parietal-occipital-lobe-syndromes-neuropsychological-approach/

23. Fierro B, Brighina F, Bisiach E. Improving Neglect by TMS. Behavioural Neurology [Internet]. 2006 Jan [cited 2025 Mar 10];17(3–4):169–76. Available from: https://onlinelibrary.wiley.com/doi/10.1155/2006/465323

24. Karlsson JL. Influence of the myopia gene on brain development. Clinical Genetics [Internet]. 1975 Nov [cited 2025 Mar 10];8(5):314–8. Available from: https://onlinelibrary.wiley.com/doi/10.1111/j.1399-0004.1975.tb01508.x

25. Saw SM, Tan SB, Fung D, Chia KS, Koh D, Tan DTH. et al. IQ and the Association with Myopia in Children. Invest Ophthalmol Vis Sci [Internet]. 2004 Sep 1 [cited 2025 Mar 10];45(9):2943. Available from: http://iovs.arvojournals.org/article.aspx?doi=10.1167/iovs.03-1296

26. Schaeffel F, Swiatczak B. Mechanisms of emmetropization and what might go wrong in myopia. Vision Res. 2024 Jul;220:108402.

27. Linke SJ, Baviera J, Munzer G, Steinberg J, Richard G, Katz T. Association between ocular dominance and spherical/astigmatic anisometropia, age, and sex: analysis of 10,264 myopic individuals. Invest Ophthalmol Vis Sci. 2011 Nov 25;52(12):9166–73.

28. Linke SJ, Druchkiv V, Steinberg J, Richard G, Katz T. Eye laterality: a comprehensive analysis in refractive surgery candidates. Acta Ophthalmol. 2013 Aug;91(5):e363–368.

29. Brasche S, Bischof W. Daily time spent indoors in German homes--baseline data for the assessment of indoor exposure of German occupants. Int J Hyg Environ Health. 2005;208(4):247–53.

30. Fomicheva ML, Zakharova MA, Chusovlyanova SV. PHYSICAL ACTIVITY LEVEL ASSESSMENT OF POPULATION GROUPS IN THE NOVOSIBIRSK REGION. Siberian Journal of Life Sciences and Agriculture. 2023;15(4):329–50.

31. Gong JF, Xie HL, Mao XJ, Zhu XB, Xie ZK, Yang HH. et al. Relevant factors of estrogen changes of myopia in adolescent females. Chin Med J (Engl). 2015 Mar 5;128(5):659–63.

32. Carney PM. Towards an Understanding of Steroid Hormone Mediated Mechanisms of Myopia Progression: A Scleral Perspective [Internet]. University of California, Berkeley; 2019 [cited 2025 Mar 12]. Available from: https://search.proquest.com/openview/ac171c0bf7517c97123076c056313c7b/1?pq-origsite=gscholar&cbl=18750&diss=y

33. Touitou Y, Reinberg A, Touitou D. Association between light at night, melatonin secretion, sleep deprivation, and the internal clock: Health impacts and mechanisms of circadian disruption. Life Sci. 2017 Mar 15;173:94–106.

34. Robeva R, Marinova E, Andonova S, Nikolaev G, Savov A, Tanev D. et al. Melatonin Receptor 1B and Corticosteroid Receptor Polymorphisms in Infertile Women with Implantation Failure and Miscarriages. Front Biosci (Landmark Ed) [Internet]. 2023 Jun 27 [cited 2025 Mar 12];28(6):122. Available from: https://www.imrpress.com/journal/FBL/28/6/10.31083/j.fbl2806122

35. Foreman J, Salim AT, Praveen A, Fonseka D, Ting DSW, Guang He M. et al. Association between digital smart device use and myopia: a systematic review and meta-analysis. Lancet Digit Health. 2021 Dec;3(12):e806–18.

36. Philipp D, Vogel M, Brandt M, Rauscher FG, Hiemisch A, Wahl S. et al. The relationship between myopia and near work, time outdoors and socioeconomic status in children and adolescents. BMC Public Health. 2022 Nov 10;22(1):2058.

37. Cumberland PM, Bountziouka V, Hammond CJ, Hysi PG, Rahi JS, UK Biobank Eye and Vision Consortium. Temporal trends in frequency, type and severity of myopia and associations with key environmental risk factors in the UK: Findings from the UK Biobank Study. PLoS One. 2022;17(1):e0260993.

